# Liquefied Petroleum Gas or Biomass for Cooking and Effects on Blood Pressure: Results from the Household Air Pollution Intervention Network (HAPIN) Trial

**DOI:** 10.1101/2025.08.15.25333780

**Authors:** Jennifer L. Peel, Howard H. Chang, Kyle Steenland, Lindsay J. Underhill, Wenlu Ye, Kalpana Balakrishnan, Anaité Díaz-Artiga, John P. McCracken, Ghislaine Rosa, Lisa M. Thompson, Vigneswari Aravindalochanan, Dana Boyd Barr, Adly Castañaza, Yunyun Chen, Marilú Chiang, Maggie L. Clark, Víctor G. Dávila-Román, Lisa de las Fuentes, Sarada Satyamoorthy Garg, Stella M. Hartinger, Steven Harvey, Shakir Hossen, Shirin Jabbarzadeh, Michael A. Johnson, Dong-Yun Kim, Miles A. Kirby, Amy E. Lovvorn, Eric D. McCollum, Lawrence H. Moulton, Alexie Mukeshimana, Krishnendu Mukhopadhyay, Luke P. Naeher, Florien Ndagijimana, Laura Nicolaou, Jean de Dieu Ntivuguruzwa, Ricardo Piedrahita, Ajay Pillarisetti, Naveen Puttaswamy, Usha Ramakrishnan, Sankar Sambandam, Suzanne Simkovich, Sheela S. Sinharoy, Gurusamy Thangavel, Lance A. Waller, Jiantong Wang, Kendra N. Williams, William Checkley, Thomas F. Clasen, Joshua P. Rosenthal, HAPIN Investigators

## Abstract

**Background:** Exposure to household air pollution from burning coal and biomass for cooking is associated with higher blood pressure and other adverse indicators of cardiovascular disease, the leading cause of death worldwide. Evidence demonstrating that switching from biomass to liquefied petroleum gas (LPG) will reduce blood pressure is limited.

**Methods:** As part of a larger trial of 3200 households, we conducted a randomized trial of 342 women aged 40 to 79 years who lived in households using biomass for cooking in rural areas of Guatemala, India, Peru, and Rwanda to assess the effects of a free LPG stove and fuel intervention. Systolic blood pressure (one of four primary outcomes) was measured once prior to randomization and up to five times after randomization over 18 months.

**Results:** A total of 418 participants were randomized (n=209 to intervention, and n=209 to control). Mean (standard deviation) systolic blood pressure at baseline was 112.7 (14.9) mmHg and 112.6 (14.7) mmHg in intervention and control participants, respectively. Adherence within the intervention arm was high. Among the 342 participants with a valid baseline and at least one valid post-randomization blood pressure measurement, mean (SD) post-randomization average systolic blood pressure was 111.5 (13.2) mmHg in the intervention arm and 111.9 (12.5) mmHg in the control arm, with an adjusted mean of difference of - 0.56 mmHg (95% confidence interval, -2.19 to 1.07, p-value = 0.50).

**Conclusions:** We did not observe evidence that an 18-month LPG stove and fuel intervention reduced blood pressure in women.

## Introduction

Non-communicable diseases, primarily cardiovascular diseases, are the foremost causes of mortality and morbidity among adults in low- and middle-income countries (LMICs), and environmental exposures are modifiable risk factors for this burden.^1^ High blood pressure is the top risk factor for premature mortality, responsible for an estimated 10.8 million premature deaths in 2019.^1^ Air pollution is the leading environmental risk factor for premature mortality and morbidity, and is responsible for an estimated 6.67 million premature deaths in 2019, with a large proportion of those deaths due to cardiovascular disease.^1^ Household air pollution resulting from inefficient combustion of solid fuels (coal and biomass) and kerosene continues to be an important source of air pollution in LMICs, where nearly 2.6 billion people use solid fuels for cooking.^2^ Over 90% of pollution-related deaths occur in LMICs.^3^

A robust body of evidence supports a causal link between air pollution, especially fine particulate matter (PM_2.5_), and cardiovascular disease; most of this evidence is from studies evaluating ambient air pollution, drawing from epidemiology, controlled human exposure studies, and toxicology, and spanning outcomes from cardiovascular mortality, hospital admissions, incident clinical disease, and subclinical measures and biomarkers. The evidence specifically from household air pollution is more limited and has focused on blood pressure as a feasible and meaningful measure of cardiovascular health. A 2021 systematic review and meta-analysis included 4 studies evaluating the impact of introducing improved biomass stoves intended to reduce air pollution.^4^ While none of the studies (3 uncontrolled pre / post evaluations, 1 randomized trial) individually reported a statistically significant effect on blood pressure, the pooled analysis indicated a beneficial effect for systolic blood pressure but not for diastolic blood pressure. The Randomized Exposure Study of Pollution Indoors and Respiratory Effects (RESPIRE) trial evaluated the impact of an improved biomass stove and reported beneficial effects on both systolic and diastolic blood pressure.^5^ More recently, trials have evaluated the impacts of non-solid fuel interventions with inconsistent results. The Cardiopulmonary and Household Air Pollution (CHAP) trial evaluated the effect of a one-year liquefied petroleum gas (LPG) intervention among 180 women aged 25-64 years in Peru and found no effect on either systolic or diastolic blood pressure, despite high adherence to the intervention and large reductions in personal exposures.^6^ Although focused on blood pressure during pregnancy, two trials have reported beneficial effects of a non-solid fuel intervention on diastolic blood pressure^7–9^ whereas our trial—the Household Air Pollution Intervention Network (HAPIN) trial— reported no effect of LPG on gestational blood pressure.^10^

The HAPIN trial was designed to evaluate exposure and health effects after the replacement of biomass cookstoves with LPG cookstoves in four low- and middle-income countries. In previous publications, we reported that the trial had high fidelity and adherence and that the intervention resulted in a substantial reduction of personal 24-hour average PM_2.5_, black carbon, and carbon monoxide exposure during the 18 months of follow-up. The other three primary outcomes of the trial (birth weight, infant pneumonia and infant stunting at 1 year of age) have been published previously. Here, we report on the fourth primary outcome, systolic blood pressure among women aged 40-79 years who resided in the same household as the recruited pregnant woman.

## Methods

### Trial design and setting

The design and methods of the HAPIN trial have been reported previously.^11–13^ HAPIN was a parallel, individually randomized trial designed to evaluate the effect of a free LPG stove and fuel intervention on four primary outcomes (birth weight, infant pneumonia, infant stunting, and systolic blood pressure in adult women). The trial included 4 research centers: Jalapa, Guatemala; Tamil Nadu, India; Puno, Peru; and Eastern Provence, Rwanda. The sites were selected based on use of biomass as the primary cooking fuel, the ability to recruit eligible participants, and minimal other sources of air pollution. We aimed to recruit 800 eligible pregnant women in each country. In a subset of households, we also recruited an additional woman aged 40 to 79 years who lived in the same household, did not currently smoke cigarettes or use other tobacco products, were not pregnant, and did not plan to move out of the current household in the next 12 months. Women who reported taking medication for blood pressure were enrolled but were not included in the primary analysis.

### Randomization and Group Assignments

Households were randomized after baseline measurements in a 1:1 ratio stratified according to study site (one site in Guatemala, two sites in India, six sites in Peru, and one site in Rwanda). Households assigned to the intervention group received a free LPG cookstove and a continuous supply of free LPG fuel through the first birthday of the child born to the pregnant woman in the same household; households in the control groups received no intervention and were anticipated to continue using primarily biomass (wood, dung, agricultural residue) for cooking fuel. Investigators not working directly with participants were blinded to assignment (except for two designated investigators who provided unblinded data to the data safety and monitoring board); participants, investigators and staff working directly with participants in the field were not blinded to assignment. In intervention households, we monitored biomass fuel use after distribution of the LPG cookstove through field worker observation, participant self-report, and with temperature sensors mounted on any biomass stoves in the house; if biomass stove use was flagged, field workers followed up to reinforce and encourage exclusive LPG use.^14^ Participants received compensation as described previously.^15^ The trial was registered in clinicaltrials.gov (Identifier NCT02944682) and approved by relevant ethics committees.

### Outcomes

Blood pressure measurements were taken 6 times during 18 months of follow-up at the time of other home visits: prior to randomization, at baseline; and post-randomization, at 24-28 weeks and 32-36 weeks gestation of the pregnant woman; and when the child was 3, 6 and 12 months of age. Resting blood pressure was measured following recommendations by the American Heart Association and the European Society of Hypertension.^16,17^ Blood pressure was measured in the right arm of seated participants by a nurse or trained field worker. Systolic and diastolic blood pressure were measured using an automatic blood pressure monitor (model HEM-907XL; Omron®) in triplicate with at least a 2-minute resting period between measurements,^11^ and average systolic (or diastolic) blood pressure was calculated. Measurements were taken after ensuring that the woman had not smoked, consumed alcohol or a caffeinated beverage, or cooked in the 30 minutes prior to the measurement.^11^ Values <70 mmHg for systolic blood pressure and <35 mmHg for diastolic blood pressure were considered implausible and removed. The average of up to three readings was calculated for each measurement time point. Systolic blood pressure was designated as one the four primary outcomes of the trial.^11^ Secondary outcomes were diastolic blood pressure, pulse pressure (systolic pressure – diastolic pressure), and mean arterial pressure ([systolic blood pressure + 2^*^diastolic pressure]/3).

### Statistical Analyses

The statistical analysis plan was published on clinicaltrials.gov in advance of the analysis. Analyses were replicated independently. Assuming the number of post-randomization measurements to be 5, an intraclass correlation of ρ=0.76, 80% power and α=0.0125 (adjusted for the four primary outcomes in the trial), and 200 women per study arm, we estimated the minimum detectable difference in mean systolic blood pressure to be 2.58 mmHg. The primary analysis excluded women who reported using blood pressure medication at enrollment or at any visit (n=41).

The primary analysis was intention-to-treat and compared the average of up to five post-randomization mean blood pressure values by study group in a linear regression model, adjusting for baseline blood pressure and randomization strata; adjusted mean differences and 95% confidence intervals for the mean differences are reported. We evaluated statistical significance of the primary outcome using a type I error rate of 0.0125 to control for family-wise type I error rate to be 0.05 under potential dependence structures among the four primary outcomes of the trial. We performed pre-specified subgroup analyses according to country, age (continuous and using median age, 51 years), and body mass index (BMI; continuous and categorical: underweight/normal, <25kg/m^2^; overweight and obese, >=25kg/m^2^).

We conducted several secondary analyses. We evaluated the change in blood pressure from baseline to the final post-randomization visit as the dependent variable without adjusting adjust for baseline blood pressure.^18^ We also conducted secondary analyses including the 41 participants taking blood pressure medication, or including participants up to the point of taking medication, for those who started taking medication after baseline (n=17).

We also conducted a repeated measures analysis, with a random intercept for individual participants, to take into account the correlation among repeated measures over time. In this analysis we compared the post-randomization blood pressure measurements between study arms, across all visits. Finally, we compared the time trend (linear slope) between intervention and control groups with a repeated measures analysis (and a random intercept for participant), coding the consecutive visits (starting at baseline) as 1, 2, 3, 4, 5, 6.

## Results

### Trial Participants

Between May 7, 2018 and February 29, 2020, a total of 418 women aged 40 to 79 years underwent randomization; 209 were assigned to the intervention group and 209 were assigned to the control group (Figure 1). Nine women were found to be ineligible after randomization (7 in the intervention group; 2 in the control group); and 41 were excluded from the primary analysis due to use of blood pressure medication at any point during the trial. At each visit, blood pressure was measured three times, and the average of the three measurements was calculated. The final sample size for the primary analysis was 342 women: additional exclusions included invalid baseline blood pressure and lack of valid post-randomization measurements. Participants who had a valid baseline blood pressure measurement and at least one valid post-randomization measurement were included. Missing data (due to drop out and missed measurement visits due to COVID-19 restrictions) was similar in intervention and control groups (Figure 1).

Baseline characteristics were similar between intervention and control groups (Table 1). The mean age at baseline was 52.0 years in the intervention group and 51.3 years in the control group; participants mostly had no formal education or did not complete primary school (80% and 78% in the intervention and control groups, respectively. Participants included in this analysis were enrolled at all four research centers (n=108 in Guatemala, n=89 in India, n=111 in Peru, n=34 in Rwanda).

**Table 1.** Characteristics of the participants at baseline.

| Characteristic | Control<br>(N=168) | Intervention<br>(N=174) |
| --- | --- | --- |
| **Age** - Mean (SD) | 51.3 (7.18) | 52.0 (7.71) |
| **Highest education completed** – n (%) |  |  |
| No formal education / primary school incomplete | 131 (78.0%) | 139 (79.9%) |
| Primary school complete / secondary school incomplete | 28 (16.7%) | 30 (17.2%) |
| Secondary school complete / vocational / some college or university | 5 (3.0%) | 2 (1.1%) |
| Missing | 4 (2.4%) | 3 (1.7%) |
| **Body Mass Index** - Mean (SD) (kg/m<sup>2</sup>) | 25.0 (5.0) | 25.3 (5.2) |
| **Minimum dietary diversity, Category (score) - n (%)** * |  |  |
| Low | 100 (59.5%) | 112 (64.4%) |
| Medium | 52 (31.0%) | 47 (27.0%) |
| High | 16 (9.5%) | 15 (8.6%) |
| **Household food insecurity, Category (score) – n (%)** ** |  |  |
| None | 84 (50.0%) | 105 (60.3%) |
| Mild | 50 (29.8%) | 54 (31.0%) |
| Moderate/Severe | 30 (17.9%) | 14 (8.0%) |
| Missing | 4 (2.4%) | 1 (0.6%) |
| **Number of people who sleep in this house** - Mean (SD) | 6.0 (2.4) | 6.0 (2.4) |
| **Someone in the household smokes** – n (%) |  |  |
| Yes | 24 (14.3%) | 21 (12.1%) |
| No | 143 (85.1%) | 153 (87.9%) |
| Missing | 1 (0.6%) | 0 (0%) |
| **Household assets owned** - n (%) |  |  |
| Color television | 93 (55.4%) | 92 (52.9%) |
| Radio | 78 (46.4%) | 87 (50.0%) |
| Mobile telephone | 153 (91.1%) | 160 (92.0%) |
| Bicycle | 46 (27.4%) | 43 (24.7%) |
| Bank account | 81 (48.2%) | 79 (45.4%) |

\*The dietary diversity score is derived from the Minimum Dietary Diversity for Women (MDD-W) questionnaire, which we adapted to cover a 30-day reference period. In the MDD-W, minimum dietary diversity is defined as consuming at least 5 of 10 food groups in the previous day. (https://www.fao.org/3/i5486e/i5486e.pdf)
\*\*Household food insecurity is measured by the Food Insecurity Experience Scale (FIES), which was applied with a 30-day reference period. In the FIES, higher scores represent increasingly severe food insecurity. (https://www.fao.org/fileadmin/templates/ess/voh/FIES_Technical_Paper_v1.1.pdf)

### Intervention Fidelity and Adherence

Fidelity and adherence were high in the HAPIN trial overall, and in the subset of households that included the additional participants described here. Of these households in which a traditional biomass stove was monitored (n=143), biomass stove use was detected on a median of 0.4% of days (Q1, Q3: 0.0%, 3.6%) for a median of 0.1 days with traditional stove use per 30 days of monitoring.

### Personal Exposure to Air Pollution

Results of the effects of the LPG intervention on average 24-hour personal exposure to PM_2.5_, black carbon, and carbon monoxide are presented elsewhere.^19,20^ Mean exposure at baseline was similar for intervention and control groups. After randomization, median 24-hour personal exposures were substantially lower in the intervention group compared to the control group. Median (25^th^ percentile, 75^th^ percentile) of the averaged PM_2.5_ measurements post-randomization was 27.8 μg/m^3^ (14.6, 39.4) in intervention households and 68.1 μg/m^3^ (35.7, 122.0) in control households.

### Primary Outcome

The mean (SD) systolic blood pressure post-randomization among the 342 women included in the analysis was 111.5 mm Hg (13.2) in the intervention group and 111.9 mmHg (12.5) in the control group (Table 2). The difference in mean blood pressure was estimated to be -0.56 mm Hg (95% confidence interval, -2.19 to 1.07, p = 0.501 > α=0.0125), adjusting for baseline systolic blood pressure and randomization strata.

**Table 2.** Adjusted mean difference in blood pressure (mm Hg) from the primary analysis, comparing intervention to control group (negative values favors intervention)

| **Outcome** | **Adjusted Mean Difference in Mean Post-Randomization Blood Pressure (95% Confidence Interval, n=342)\*** | **Adjusted Mean Difference in Change from Baseline to Final Post-Randomization Visit (95% Confidence Interval, n=223)\*\*** |
| --- | --- | --- |
| Systolic Blood Pressure - mmHg | -0.56 (-2.19, 1.07) | 0.20 (-3.03, 3.42) |
| Diastolic Blood Pressure - mmHg | -1.21 (-2.41, -0.01) | -1.90 (-4.52, 0.71) |
| Pulse Pressure - mmHg | 0.67 (-0.45, 1.79) | 2.10 (-0.49, 4.68) |
| Mean Arterial Pressure - mmHg | -1.02 (-2.27, 0.23) | -1.20 (-3.76, 1.35) |

\*Adjusted mean differences of mean post-randomization blood pressure, comparing intervention to control groups, adjusting for centered mean baseline blood pressure and randomization strata
\*\*Adjusted mean differences in change from baseline to final post-randomization visit, comparing intervention to control groups, adjusting for centered mean baseline blood pressure and randomization strata

### Subgroup Analyses and Secondary Outcomes

In subgroup analyses of systolic blood pressure by study site, we observed stronger but imprecise effects in India, Peru, and Rwanda and a weaker effect in the opposite direction in Guatemala (Figure 2). We observed a somewhat stronger effect in older women (>=51 years) than in younger women (40 - <51 years) (Figure 2). For subgroup analyses by body mass index (BMI), we observed an effect favoring the intervention among those in the normal and underweight categories and an effect favoring the control group among those who were in the obese and overweight categories, although confidence intervals were wide (Figure 2). Results for diastolic blood pressure provided some evidence of a benefit for this secondary outcome (Table 2; Supplemental Figures 1-3). For the other secondary outcomes (pulse pressure, mean arterial pressure) trial-wide and subgroup results were generally similar to those observed for the primary outcome (Table 2; Supplemental Figures 1-3).

### Secondary Analyses

In secondary analyses comparing the change in blood pressure from baseline to the final post-randomization visit (n=248) in intervention to control groups (adjusting for randomization strata), we did not observe an effect of the intervention on any of the blood pressure outcomes; the 95% confidence intervals were relatively wide (Table 2).

In secondary analyses, including measurements from women taking blood pressure medication after randomization showed little change in results (the difference in mean post-randomization SBP was 0.23 mmHg, 95% CI: -1.52, 1.98), nor did including only measurements prior to when they began taking medication (0.08, 95% CI: -1.68, 1.84) (Supplemental Table 1). Analogous results for other endpoints (diastolic blood pressure, mean arterial pressure, and pulse pressure) also showed little change after including women taking blood pressure medication.

Secondary analyses using repeated measures to evaluate the post-randomization SBP, while taking into account differing number of repeated measures and including a random intercept for women, also did not show any difference between study arms (average SPB for intervention vs control was -0.01 (95% CI: - 2.52, 2.50)). Again, there were also no marked differences between arms for other outcomes (diastolic blood pressure, mean arterial pressure, and pulse pressure).

Finally, the time trends (slopes) for both intervention and control groups over time also showed little difference between study arms, using a repeated measures analysis, with consecutive time points coded 1 through 6. Both groups showed decreased SBP over follow-up. The slope for the intervention group was - 0.41 (95% CI: -0.77, -0.05) while for the control group it was -0.40 (95% CI: -0.76, -0.04); the adjusted difference in slope between arms was -0.01 (95% CI: -0.52, 0.49) (Supplemental Table 1). Analogous analyses for other outcomes also showed no differences.

## Discussion

Despite high levels of intervention compliance and substantial contrasts in exposure to measured pollutants, we did not find a difference in systolic blood pressure or secondary blood pressure outcomes in women residing in households using LPG cooking fuel for 18 months compared to those residing in households relying on biomass fuels. Our findings are consistent with results reported from the CHAP trial in Peru that reported a difference of 0.7 mmHg (95% CI: -2.1, 3.4) for systolic blood pressure in intervention compared to control participants.^6^ They are also consistent with a trial of an improved biomass cookstove in India, though that intervention did not achieve reductions in measured kitchen concentrations.^21^ The suggestive beneficial effect on diastolic blood pressure, although a secondary outcome, is consistent with results from several other studies,^5,7^ as well as the results for gestational blood pressure from the HAPIN trial.^10^

Our results should be considered in the context of several factors. Although we achieved substantial reductions in personal exposure, even further reductions may need to be achieved to observe measurable benefits to health over the time period considered. For many health outcomes, there is no evidence of a threshold or a level below which there are no adverse effects on health. A controlled human exposure study reported short increases in blood pressure after exposure to emissions from LPG stoves compared to clean air.^22^ Further, increases in risk may actually be greater at lower concentrations, given the modeled shape of exposure-response curves for several health outcomes.^1^ It is also possible that blood pressure is not reversable in this context after a lifetime of elevated exposures; interventions may need to occur earlier in life to reduce cumulative lifetime exposure, although evidence from smoking cessation and from controlled human exposure studies of air pollution suggests an acute reduction in blood pressure after removing exposure.^22^ Error in blood pressure measurements could theoretically lead to an attenuation of observed effect; however, our standardized protocols and training and use of an automated device would likely diminish this concern. Although we did have missing data and loss to follow-up, the missing data was balanced between study arms and did not likely result in a selection bias; however, the sample size may have limited statistical power to detect differences in this scenario. The randomization also appeared to result in balanced study groups, minimizing concern of confounding. Finally, diastolic blood pressure may be more sensitive to the effect of the LPG intervention than systolic blood pressure, consistent with some, but not all, previous randomized trials.^5–8^

Despite inclusion of multiple geographic regions, absence of other important sources of background air pollution, and achievement of a relatively low exposure to particulate matter in the intervention group, we did not observe benefits of the 18-month LPG intervention on systolic blood pressure, an important risk factor globally for premature mortality and disability. While these results do not exclude the possibility that LPG or other clean energy solutions, such as electricity, could have other health benefits, our results do not support the use of LPG interventions to improve systolic blood pressure under these conditions.

## Data Availability

All data produced in the present work are available upon reasonable request to the authors

## Acknowledgements

A multidisciplinary, independent Data and Safety Monitoring Board (DSMB) appointed by the National Heart, Lung, and Blood Institute (NHLBI) monitored the quality of the data and protected the safety of patients enrolled in the HAPIN trial. The DSMB consisted of: Catherine Karr (Chair), Nancy R. Cook, Stephen Hecht, Joseph Millum, Nalini Sathiakumar (deceased), Paul K. Whelton, and Gail Weinmann and Thomas Croxton (Executive Secretaries). Program Coordination: Gail Rodgers, Bill & Melinda Gates Foundation; Claudia L. Thompson, National Institute of Environmental Health Sciences; Mark J. Parascandola, National Cancer Institute; Marion Koso-Thomas, Eunice Kennedy Shriver National Institute of Child Health and Human Development; Joshua P. Rosenthal, Fogarty International Center; Concepcion R. Nierras, NIH Office of Strategic Coordination – The Common Fund; Katherine Kavounis, Dong-Yun Kim, Barry S. Schmetter (deceased), and Antonello Punturieri, NHLBI. This research represents the NIH’s contribution to the Global Alliance for Chronic Diseases (GACD) coordinated call for research on prevention and management of chronic lung diseases for 2016.

The investigators would like to thank the Drs. Patrick Breysse, Donna Spiegelman, and Joel Kaufman (members of the advisory committee) for their valuable insight and guidance throughout the implementation of the trial. We also wish to acknowledge all the research staff and study participants for their dedication to and participation in this important trial.

## Ethics Approvals

The study protocol was reviewed and approved by institutional review boards (IRBs) or Ethics Committees at Emory University (00089799), Johns Hopkins University (00007403), Sri Ramachandra Institute of Higher Education and Research (IEC-NI/16/JUL/54/49) and the Indian Council of Medical Research – Health Ministry Screening Committee (5/8/4-30/(Env)/Indo-US/2016-NCD-I), Universidad del Valle de Guatemala (146-08-2016) and Guatemalan Ministry of Health National Ethics Committee (11-2016), Asociación Benefica PRISMA-Peru (CE2981.17; CE2008.18; CE0028.20; CE0291.21), the London School of Hygiene and Tropical Medicine (11664) and the Rwandan National Ethics Committee (853/RNEC/2016; 317/2017; 357/2018; 194/2019; 929/2020; 64/2021), and Washington University in St. Louis (201611159). The study has been registered with clinicaltrials.gov (Identifier NCT02944682).

## References

1. Murray CJL, Aravkin AY, Zheng P, et al. Global burden of 87 risk factors in 204 countries and territories, 1990–2019: a systematic analysis for the Global Burden of Disease Study 2019. The Lancet 2020;396(10258):1223–49.

2. Stoner O, Lewis J, Martínez IL, Gumy S, Economou T, Adair-Rohani H. Household cooking fuel estimates at global and country level for 1990 to 2030. Nature Communications 2021;12(1):5793.

3. Fuller R, Landrigan PJ, Balakrishnan K, et al. Pollution and health: a progress update. The Lancet Planetary Health 2022;6(6):e535–47.

4. Kumar N, Phillip E, Cooper H, et al. Do improved biomass cookstove interventions improve indoor air quality and blood pressure? A systematic review and meta-analysis. Environmental Pollution 2021;290:117997.

5. McCracken JP, Smith KR, Diaz A, Mittleman MA, Schwartz J. Chimney stove intervention to reduce long-term wood smoke exposure lowers blood pressure among Guatemalan women. Environmental health perspectives 2007;115(7):996–1001.

6. Checkley W, Williams KN, Kephart JL, et al. Effects of a Household Air Pollution Intervention with Liquefied Petroleum Gas on Cardiopulmonary Outcomes in Peru. A Randomized Controlled Trial. Am J Respir Crit Care Med 2021;203(11):1386–97.

7. Alexander D, Northcross A, Wilson N, et al. Randomized Controlled Ethanol Cookstove Intervention and Blood Pressure in Pregnant Nigerian Women. American Journal of Respiratory and Critical Care Medicine 2017;195(12):1629:1639.

8. Quinn AK, Ae-Ngibise KA, Jack DW, et al. Association of Carbon Monoxide exposure with blood pressure among pregnant women in rural Ghana: Evidence from GRAPHS. International journal of hygiene and environmental health 2016;219(2):176–83.

9. Quinn AK, Ae-Ngibise KA, Kinney PL, et al. Ambulatory monitoring demonstrates an acute association between cookstove-related carbon monoxide and blood pressure in a Ghanaian cohort. Environmental Health 2017;16(1):76.

10. Ye W, Steenland K, Quinn A, et al. Effects of a Liquefied Petroleum Gas Stove Intervention on Gestational Blood Pressure: Intention-to-Treat and Exposure-Response Findings From the HAPIN Trial. Hypertension 2022;79(8):1887–98.

11. Clasen T, Checkley W, Peel JL, et al. Design and Rationale of the HAPIN Study: A Multicountry Randomized Controlled Trial to Assess the Effect of Liquefied Petroleum Gas Stove and Continuous Fuel Distribution. Environ Health Perspect 2020;128(4):047008.

12. Barr Dana Boyd, Puttaswamy Naveen, Jaacks Lindsay M., et al. Design and Rationale of the Biomarker Center of the Household Air Pollution Intervention Network (HAPIN) Trial. Environmental Health Perspectives 2020;128(4):047010.

13. Johnson MA, Steenland K, Piedrahita R, et al. Air Pollutant Exposure and Stove Use Assessment Methods for the Household Air Pollution Intervention Network (HAPIN) Trial. Environ Health Perspect 2020;128(4):47009.

14. Williams KN, Thompson LM, Sakas Z, et al. Designing a comprehensive behaviour change intervention to promote and monitor exclusive use of liquefied petroleum gas stoves for the Household Air Pollution Intervention Network (HAPIN) trial. BMJ Open 2020;10(9):e037761.

15. Quinn AK, Williams K, Thompson LM, et al. Compensating control participants when the intervention is of significant value: experience in Guatemala, India, Peru and Rwanda. BMJ Glob Health 2019;4(4):e001567.

16. Parati G, Stergiou G, O’Brien E, et al. European Society of Hypertension practice guidelines for ambulatory blood pressure monitoring. J Hypertens 2014;32(7):1359–66.

17. Pickering TG, Hall JE, Appel LJ, et al. Recommendations for Blood Pressure Measurement in Humans and Experimental Animals. Hypertension 2005;45(1):142–61.

18. Glymour MM, Weuve J, Berkman LF, Kawachi I, Robins JM. When Is Baseline Adjustment Useful in Analyses of Change? An Example with Education and Cognitive Change. American Journal of Epidemiology 2005;162(3):267–78.

19. Johnson M, Pillarisetti A, Piedrahita R, et al. Exposure Contrasts of Pregnant Women during the Household Air Pollution Intervention Network Randomized Controlled Trial. Environ Health Perspect 2022;130(9):97005.

20. Ye W, Campbell D, Johnson M, et al. Exposure Contrasts of Women Aged 40–79 Years during the Household Air Pollution Intervention Network Randomized Controlled Trial. Environ Sci Technol [Internet] 2024 [cited 2024 Dec 31];Available from: 10.1021/acs.est.4c06337

21. Aung TW, Baumgartner J, Jain G, et al. Effect on blood pressure and eye health symptoms in a climate-financed randomized cookstove intervention study in rural India. Environmental Research 2018;166:658–67.

22. Fedak KM, Good N, Walker ES, et al. Acute Effects on Blood Pressure Following Controlled Exposure to Cookstove Air Pollution in the STOVES Study. Journal of the American Heart Association 2019;8(14):e012246.

