## Supplemental tables S1 and S2 for "Liquefied Petroleum Gas or Biomass for Cooking and Effects on Blood Pressure: Results from the Household Air Pollution Intervention Network (HAPIN) Trial": HAPIN BP supplemental tables S1_S2.pdf

**Table S1. Measured effect of the HAPIN intervention on blood pressure using primary and secondary analytical models across four participant subsets with varying levels of medication use.** All models evaluate differences in blood pressure between study arms. Model 1 utilizes mean blood pressure after randomization as the outcome and adjusts for centered mean baseline blood pressure and randomization strata. Model 2 uses the change in blood pressure from baseline to the final post-randomization visit as the outcome and adjusts for randomization strata but not baseline blood pressure. Model 3 utilizes a repeated measures analysis incorporating a random effect for women to compare average post-randomization blood pressure between study arms. Model 4 compares the time trend (slope) of blood pressure changes between study arms using a repeated measures analysis with a random intercept for women, coding consecutive visits numerically from baseline onward. All models were applied to three participant subsets: The "after" subset excluded measurements at and after first report of medication usage; the "BL" subset excluded all participants reporting medication use at baseline only; and the "Any" subset excluded participants who reported medication use at any timepoint. The model and subsets used in the primary analysis are indicated by an asterisk.

| | | Model 1* Post-Randomization Mean Difference | | | Model 2 Change (Baseline to 18-Months Post-Randomization) | | | Model 3 Repeated measures + Random Effects | | | Model 4 Repeated measures + Slopes Comparison | | |
|---|---|---|---|---|---|---|---|---|---|---|---|---|---|
| | Exclusion | n | Est. | 95% CI | n | Est. | 95% CI | n | Est. | 95% CI | n | Est. | 95% CI |
| SBP | After | 352 | 0.08 | (-1.68, 1.84) | NA | NA | NA | 1262 | 0.83 | (-1.86, 3.52) | 1621 | 0.02 | (-0.49, 0.53) |
| | BL | 359 | 0.23 | (-1.52, 1.98) | 248 | -0.11 | (-3.46, 3.24) | 1304 | 0.82 | (-2.03, 3.66) | 1663 | 0.07 | (-0.45, 0.6) |
| | Any* | 342 | -0.56 | (-2.19, 1.07) | 248 | 0.20 | (-3.03, 3.42) | 1244 | -0.01 | (-2.52, 2.5) | 1586 | -0.01 | (-0.52, 0.49) |
| DBP | After | 352 | -0.85 | (-2.1, 0.4) | NA | NA | NA | 1262 | 0.13 | (-1.54, 1.8) | 1621 | -0.30 | (-0.67, 0.08) |
| | BL | 359 | -0.91 | (-2.15, 0.33) | 248 | -1.82 | (-4.38, 0.75) | 1304 | 0.09 | (-1.64, 1.82) | 1663 | -0.23 | (-0.6, 0.14) |
| | Any* | 342 | -1.21 | (-2.41, -0.01) | 248 | -1.90 | (-4.52, 0.71) | 1244 | -0.14 | (-1.78, 1.49) | 1586 | -0.32 | (-0.69, 0.06) |
| PP | After | 352 | 1.04 | (-0.14, 2.22) | NA | NA | NA | 1262 | 0.68 | (-0.99, 2.34) | 1621 | 0.33 | (-0.05, 0.7) |
| | BL | 359 | 1.20 | (0.03, 2.37) | 248 | 1.71 | (-0.98, 4.4) | 1304 | 0.74 | (-1, 2.47) | 1663 | 0.31 | (-0.08, 0.71) |
| | Any* | 342 | 0.67 | (-0.45, 1.79) | 248 | 2.10 | (-0.49, 4.68) | 1244 | 0.16 | (-1.4, 1.71) | 1586 | 0.32 | (-0.05, 0.69) |
| MAP | After | 352 | -0.74 | (-2, 0.52) | NA | NA | NA | 1262 | 0.35 | (-1.56, 2.26) | 1621 | -0.20 | (-0.58, 0.19) |
| | BL | 359 | -0.56 | (-1.88, 0.76) | 248 | -1.25 | (-3.8, 1.31) | 1304 | 0.31 | (-1.7, 2.32) | 1663 | -0.14 | (-0.52, 0.25) |
| | Any* | 342 | -1.02 | (-2.27, 0.23) | 248 | -1.20 | (-3.76, 1.35) | 1244 | -0.12 | (-1.95, 1.72) | 1586 | -0.22 | (-0.6, 0.16) |

**Table S2: Measured effect of the HAPIN intervention on blood pressure over time within control and intervention arms.** Estimates represent the change in mean blood pressure over time (slope) within each study arm, using a repeated measures analysis with a random intercept for women, with visits coded sequentially from baseline onward (i.e., Model 4 from Table S1, stratified by study arm). All models were applied to the three participant subsets defined in Table S1.

| | | Model 4 (Control) | | | Model 4 (Intervention) | | |
|---|---|---|---|---|---|---|---|
| | Exclusion | n | Est. | 95% CI | n | Est. | 95% CI |
| SBP | After | 795 | -0.48 | (-0.84, -0.12) | 826 | -0.47 | (-0.83, -0.11) |
| SBP | BL | 815 | -0.56 | (-0.94, -0.19) | 848 | -0.49 | (-0.86, -0.12) |
| SBP | Any | 783 | -0.40 | (-0.76, -0.04) | 803 | -0.41 | (-0.77, -0.05) |
| DBP | After | 795 | 0.03 | (-0.23, 0.29) | 826 | -0.27 | (-0.53, -0.01) |
| DBP | BL | 815 | 0.01 | (-0.25, 0.28) | 848 | -0.22 | (-0.48, 0.04) |
| DBP | Any | 783 | 0.07 | (-0.2, 0.33) | 803 | -0.25 | (-0.51, 0.01) |
| pulse | After | 795 | -0.53 | (-0.8, -0.27) | 826 | -0.21 | (-0.47, 0.06) |
| pulse | BL | 815 | -0.58 | (-0.85, -0.3) | 848 | -0.26 | (-0.54, 0.01) |
| pulse | Any | 783 | -0.46 | (-0.73, -0.2) | 803 | -0.14 | (-0.41, 0.12) |
| MAP | After | 795 | -0.14 | (-0.41, 0.13) | 826 | -0.34 | (-0.61, -0.07) |
| MAP | BL | 815 | -0.18 | (-0.45, 0.1) | 848 | -0.31 | (-0.59, -0.04) |
| MAP | Any | 783 | -0.09 | (-0.36, 0.18) | 803 | -0.31 | (-0.58, -0.04) |
